# Chronotype is Associated with Sleep Quality in Older Adults

**DOI:** 10.1101/2023.09.04.23294997

**Authors:** Scott C. Sauers, Cristina D. Toedebusch, Rachel Richardson, Adam P. Spira, John C. Morris, David M. Holtzman, Brendan P. Lucey

## Abstract

**Introduction:** Disrupted sleep is common in individuals with Alzheimer’s disease (AD) and may be a marker for AD risk. The timing of sleep or chronotype affects sleep-wake activity and is also associated with AD, but little is known about links between sleep and chronotype in older adults. In this study, we tested if different measures of sleep and chronotype are associated among older adults even after adjusting for multiple potentially confounding variables.

**Methods:** Participants (N=243) with a mean age of 74 underwent standardized cognitive assessments, measurement of CSF AD biomarkers, and sleep monitoring via single-channel EEG, actigraphy, and self-reported sleep logs. Chronotype was defined as the midpoint of sleep measured by actigraphy.

**Results:** Later mid-point of sleep (i.e., late chronotype) was associated with African American race and greater night-to-night variability in the sleep mid-point. After controlling for age, race, sex, cognitive status, AD biomarkers, and sleep disorders, a later mid-point of sleep was associated with longer rapid eye movement (REM) onset latency, decreased REM sleep time, lower sleep efficiency, increased sleep onset latency, and more awakenings at night. Late chronotype was also associated with increased <2 Hz non-REM slow-wave activity.

**Conclusions:** To identify individuals at risk for cognitive impairment before symptoms onset, non-invasive *in vivo* markers of brain function, such as sleep, are needed to track both future risk of cognitive impairment and response to interventions. Chronotype is a potential modifiable AD risk factor and should also be taken into account when using sleep as a marker for AD risk.

## INTRODUCTION

Alzheimer’s disease (AD) is a progressive neurodegenerative disorder characterized neuropathologically by insoluble extracellular plaques of amyloid-β (Aβ) and intracellular neurofibrillary tangles of hyperphosphorylated tau resulting in cognitive decline, dementia, and death.^1^ AD is a growing public health crisis with the global prevalence of AD and other dementias expected to increase from 57 million cases as of 2019 to 153 million cases in 2050.^2^ Amyloid pathology emerges ∼15-20 years before synaptic dysfunction, neuronal loss, and subsequent cognitive decline.^3^ Although there are many markers for AD pathology including the soluble cerebrospinal fluid (CSF) concentration of Aβ42,^4^ the CSF and plasma Aβ42/40 ratio,^5,6^ the levels of CSF and plasma p-tau species,^7,8^ and radiotracers that bind to Aβ and tau via positron emission tomography (PET),^9–11^ a major goal of the field is to develop AD biomarkers of brain function to identify individuals who are at risk for cognitive impairment before symptom onset. Sleep is a potential non-invasive *in vivo* marker of brain function that could be followed to track future risk of cognitive impairment as well as response to interventions such as anti-amyloid monoclonal antibodies.

Multiple measures of sleep and sleep disorders have been associated with an increased risk of AD or cognitive decline including poor self-reported sleep quality,^12^ short or long sleep durations,^13^ fragmented sleep,^14^ decreased sleep efficiency,^15^ increased wake after sleep onset,^16^ increased sleep onset latency,^17^ increased rapid eye movement (REM) sleep onset latency,^18^ time spent in different sleep stages,^19,20^ sleep-disordered breathing,^21^ and periodic limb movement during sleep.^22^ Sleep and AD are hypothesized to have a bi-directional relationship with disrupted sleep contributing to the development of AD and AD pathology resulting in sleep disturbances.^23,24^ Disrupted circadian rhythms are also associated with increased risk of AD and future risk of cognitive impairment. For instance, lower rest-activity rhythm amplitude is linked to AD and dementia.^25^ Neurodegenerative diseases such as AD disrupt the circadian system leading to unpredictable rhythms and fragmented sleep. AD has been associated with delayed circadian phase.^26,27^ Abnormalities in the phase of circadian rhythms and abnormalities in behavioral chronotype are also associated with risk or severity _of AD._25,28

Chronotype is the body’s natural time to sleep and regulates timing of sleep and wake activity.^29^ The midpoint of sleep or the midpoint of time in bed may be measured by actigraphy or sleep logs, is analogous to the trough of the circadian rest-activity rhythm, and has been proposed as a measure of chronotype.^30–34^ Chronotype is associated with a wide variety of diseases. In general, individuals with later chronotypes are at risk for worse health outcomes, including increased risk for dementia.^27,35,36^ Mendelian randomization analyses suggest a causal role of late chronotype and increasing digestive tract cancer,^37^ prostate cancer,^38^ poor mental health,^39^ and educational attainment,^40,41^ as well as a role of cognitive function in causing later chronotype.^42^ In the UK Biobank dataset, chronotype has a positive genetic correlation with educational attainment and intelligence, and a negative genetic correlation with time spent outdoors, physical activity, and daytime napping.^43^ Individuals with late chronotypes are more at risk for spinal, gastrointestinal, respiratory, and cardiovascular diseases, as well as depression, anxiety, personality disorders, substance use disorders, infertility, obesity, diabetes, insomnia, sleep apnea, and all-cause mortality.^44^

The potential to use sleep measures as a marker for AD risk is likely to be affected by timing of sleep or chronotype given the overlapping relationships between sleep, circadian rhythms, and AD. In this study, we tested if different measures of sleep and chronotype are related even after adjusting for multiple confounding variables such as age, sex, race, AD biomarkers, and cognitive status. We hypothesized that poorer measures of sleep quality (e.g., lower sleep efficiency) will be associated with late chronotype. If sleep measures are affected by chronotype, then chronotype may need to be taken into account when using sleep as a marker for AD risk. Chronotype may be a partial mediator or confound for a sleep measurement, which may lead to a spurious association with an outcome (e.g., AD pathology or cognitive deficit) if left unaccounted for. Further, this will also support chronotype as a potential modifiable AD risk factor.

## METHODS

### Participants

Data were gathered from 388 community-living participants enrolled in ongoing longitudinal studies at the Knight Alzheimer Disease Research Center (ADRC), Washington University in St. Louis. All individuals participating in Knight ADRC studies undergo annual standardized clinical and cognitive assessments. The Clinical Dementia Rating® (CDR®) was used to determine if participants were cognitively unimpaired (CDR 0), or mildly impaired (CDR 0.5). This study was approved by the Washington University in St. Louis Institutional Review Board. Each participant provided signed informed consent and was compensated for their participation.

### Sleep Monitoring

Sleep was recorded longitudinally at home for up to 6 nights using self-reported sleep logs, actigraphy (Actiwatch2, Philips Respironics) and a single-channel EEG device worn on the forehead (Sleep Profiler, Advanced Brain Monitoring). Sleep parameters were determined for the single-channel EEG, actigraphy, and sleep logs. Sleep-disordered breathing and periodic leg movements were measured using a home sleep apnea test (HSAT) device (Alice PDx, Philips Respironics), a device that was found to have 96.4% agreement with simultaneously recorded in-laboratory polysomnography.^45^

#### Single-Channel EEG

Average total sleep time, time in non-REM (NREM) sleep stages 2 and 3 (time in NREM), time in REM sleep, sleep efficiency, and NREM slow wave activity (SWA) were measured as previously described.^46^ Sleep efficiency was calculated based on the lights off and lights on times for the single-channel EEG studies and which were corroborated with actigraphy and sleep logs. Single-channel EEG sleep studies were visually scored by registered polysomnographic technologists using criteria adapted from the standard American Academy of Sleep Medicine (AASM) criteria.^46^ Nights were excluded if >10% of the recording was artifactual and if the bed and rise times did not match the sleep log and/or actigraphy. All participants needed at least 2 nights of single-channel EEG monitoring that met these criteria to be included in the analysis. Time in NREM sleep stages 2 and 3 were combined, as the combined metric has a higher level of agreement with polysomnography.^20^

NREM SWA was calculated for each single-channel EEG study using MATLAB (MathWorks, Natick, MA), and the average NREM SWA in different frequency bins were used in the analysis. As previously described, a band-pass (two-way least-squares finite impulse response) filter between 0.5 and 40 Hz was applied to the single-channel EEG data. Spectral analysis was performed in consecutive 5-s epochs (Welch method, Hamming window, no overlap).^20^ SWA power was calculated by averaging the power in the frequency bins of 0.5–1.0 Hz, 1.0-2.0 Hz, 2.0-3.0 Hz, and 3.0-4.0 Hz. To semi-automatically remove artifactual epochs, power in the 20–30 Hz and 0.5–4.5 Hz bands for each electrode across all epochs of a recording were displayed. The operator (B.P.L.) then selected a threshold between the 95 and 99.5% threshold of power to remove artifactual epochs.

#### Actigraphy

Actigraphy is commonly used to assess sleep and is validated against polysomnography.^47^ Start and end times were set using a standardized protocol involving event marker button presses and sleep logs as previously described.^48^ Actigraphic sleep parameters were calculated using Actiware 6.0 (Philips Respironics) and included: total sleep time, sleep efficiency, sleep onset latency, wake after sleep onset, and number of awakenings.

#### Sleep Logs

Participants completed a sleep log for all nights that the single-channel EEG, actigraphy, and the HSAT were worn. Self-reported sleep parameters included total sleep time, sleep onset latency, and the number of awakenings during the night.

#### Home Sleep Apnea Test

Participants were monitored at home for one night to measure sleep-disordered breathing and periodic leg movements. The Alice PDx is a type III HSAT device that monitors oxygen saturation (SpO2) and pulse rate from an oximeter finger probe, nasal pressure-based airflow monitor and thermistor, thoracic and abdominal effort via inductance plethysmography, bilateral electromyography sensors over the anterior tibialis muscles, and body position. Participants pressed the event button monitor at lights off and lights on. Bed and rise times were also confirmed with sleep logs and actigraphy as previously described.^48^ In the morning, participants checked the “good study” indicator on the device to confirm a minimum of 4 hours of recording. A minimum of 4 hours of artifact-free recording was obtained for all participants and participants not meeting this criterion were asked to repeat monitoring. Respiratory events and periodic leg movements were scored by registered polysomnographic technologists using AASM criteria^49^ and were reviewed by a board-certified sleep medicine physician (B.P.L.). The criteria for scoring hypopneas was a 4% decrease in oxygen saturation. The calculation of the Apnea-Hypopnea Index (AHI) and Periodic Leg Movement Index (PLMI) were calculated per hour of monitoring time for each participant. Participants using PAP therapy or dental devices were asked to use them as usual during the HSAT.

### Cerebrospinal fluid

CSF was collected under a standardized protocol.^50^ After overnight fasting, participants underwent a lumbar puncture at 8 AM and 20-30 ml of CSF was collected by gravity drip into a 50-ml conical tube using a 22-gauge atraumatic Sprotte spinal needle. The conical tube was then gently inverted to disrupt potential gradient effects and centrifuged at low speed to pellet any cellular debris. Samples were aliquoted (500 μl) in polypropylene tubes and stored at –80°C until analysis. CSF Aβ40 and Aβ42, total tau (t-tau), and phosphorylated tau-181 were measured as previously described using an automated electrochemiluminescence immunoassay (Lumipulse, Fujirebio).^51^

### Chronotype

A participant’s chronotype was defined as the average midpoint of the participant’s sleep as measured by actigraphy. Actigraphy has previously been used to determine the midpoint of sleep.^32,33^ The midpoint of sleep was calculated for individual nights based on bedtimes and risetimes, and then averaged for each participant. Midnight was set at “0” and times before midnight were negative (e.g., 11 PM = –1) and times after midnight were positive (e.g., 1 AM = +1). Participants were divided into equal groups of “early” and “late” chronotype depending on if they were before or after the average midpoint of sleep for the sample (3:04 AM). Forty-three participants also completed the Munich Chronotype Questionnaire (MCTQ)^52^ that collects information on sleep-wake schedules during work and free days. The MCTQ is used to determine the midpoint of sleep and circadian phase. Comparison of the midpoint of sleep determined by actigraphy and the MCTQ found a high correlation (Supplementary Figure 1: r=0.823; p=1.278×10^-11^). Chronotype variability was also calculated as the standard deviation of an individual’s chronotype across multiple nights of monitoring. Higher chronotype variability indicates greater night-to-night differences in the midpoint of sleep.

### Statistical analysis

Differences between chronotype groups were tested using Welch’s 2-tailed t-test, or Chi-square if the data were categorical. Linear regression models tested if chronotype predicted different sleep variables after adjusting for age, sex, race, CDR, APOE4 status, CSF Aβ42/40, AHI, and PLMI. Only participants with each of these variables were included in the main analyses. Coding for dichotomous variables in the regression model is as follows: 1 = male, 2 = female; 0 = non-African American, 1 = African American (race determined by self-report); CDR negative (a score of 0) = 1, CDR positive (a non-zero score) = 2. All analyses were performed using R. Statistical significance was set at p = 0.05.

### Data Availability

Data to support the findings of this study are available from the corresponding author upon reasonable request. All code associated with this analysis is freely available from the corresponding author upon reasonable request.

## RESULTS

### Participant Characteristics

Demographic and sleep parameter differences between late and early chronotype groups are shown in Tables 1, 2 and 3. Sex and age were not significantly different between early and late chronotype groups. African Americans were significantly more likely to have a late chronotype than non-African Americans and this relationship remained significant (p=0.007) after including participants who were missing other data and were not included in analyses using the fully adjusted model (totaling N=58 African Americans, N=293 non-African Americans). The distribution of chronotype by race, including these additional participants, is shown in Figure 1. Chronotype variability was also significantly higher in the late chronotype group.

**Figure 1:**
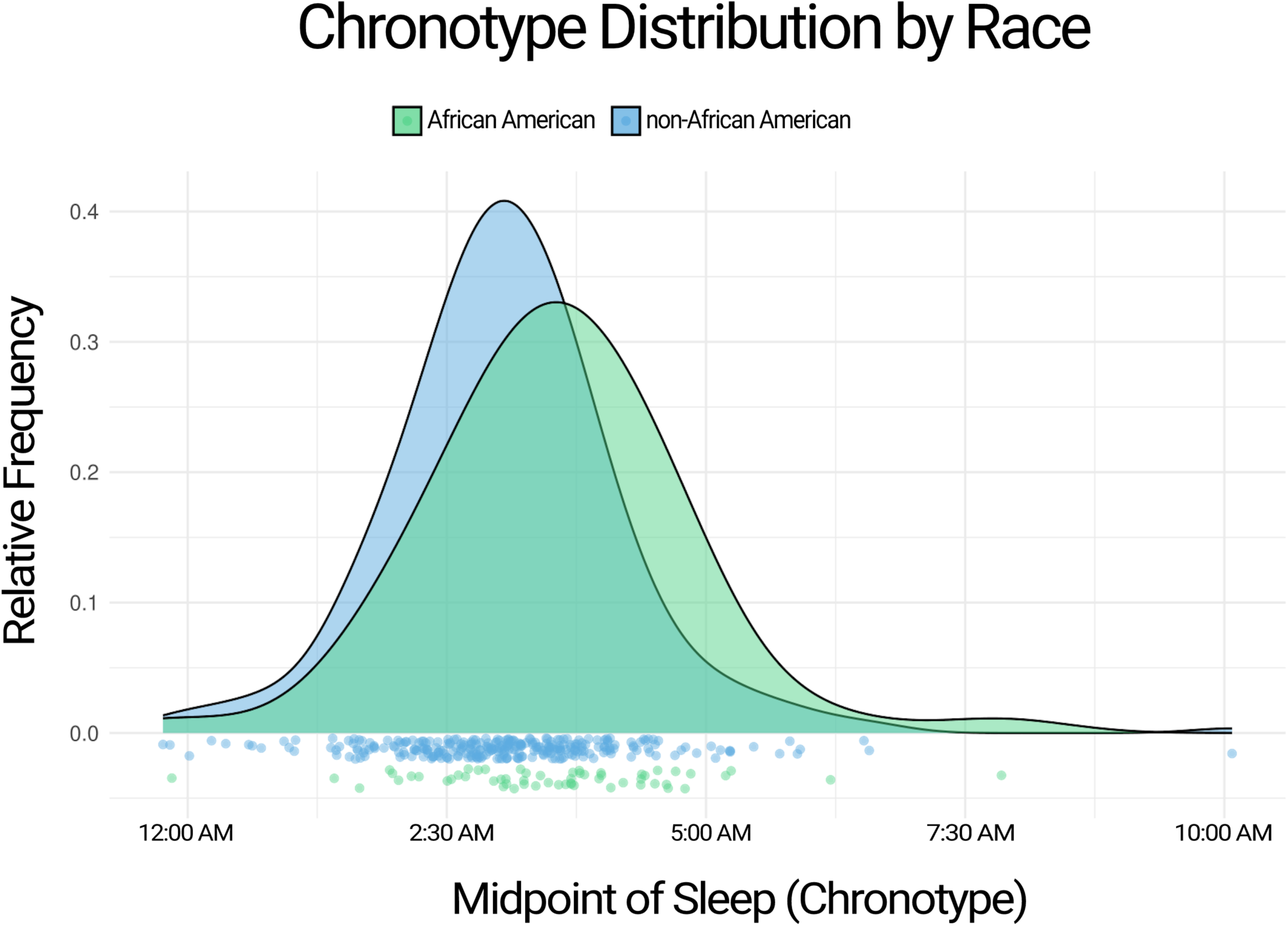
Distribution of midpoint of sleep by race. African Americans had a later midpoint of sleep (chronotype) than non-African Americans. Green = African American. Blue = non-African American.

**Table 1:** Participant Characteristics.

| **Table 1: Participant Characteristics** |  |  |  |
| --- | --- | --- | --- |
|  | **Early chronotype** | **Late chronotype** | **p-value** |
|  | Midpoint of sleep before 3:04 AM<br>(n = 122) | Midpoint of sleep after 3:04 AM<br>(n = 121) |  |
| Age at sleep study, years, mean (SD) | 73.785 (4.996) | 73.589 (5.536) | 0.7717 |
| Sex, number of females (%) | 59 (48.361%) | 68 (56.198%) | 0.2213 |
| Race, number of African Americans, n (%) | 7 (5.738%) | 17 (14.05%) | **0.0299** |
| Clinical Dementia Rating, n CDR+ (% = 0.5) | 25 (20.492%) | 22 (18.182%) | 0.6485 |
| APOE ε4 status: n APOE ε4+ (%) | 45 (36.885%) | 42 (34.711%) | 0.724 |
| CSF Aβ42/Aβ40 ratio Mean (SD) and then t-test | 0.0687 (0.0233) | 0.0734 (0.0226) | 0.1754 |
| Amyloid pathology, number Aβ+ (% CSF Aβ42/Aβ40 < 0.0673) | 62 (50.82%) | 51 (42.149%) | 0.1085 |
| AHI, number of respiratory events/h of monitoring time (%) |  |  | 0.1798 |
| None (<5) | 47 (38.525%) | 60 (49.587%) |  |
| Mild (5–<15) | 49 (40.164%) | 40 (33.058%) |  |
| Moderate (15–<30) | 20 (16.393%) | 18 (14.876%) |  |
| Severe (≥30) | 6 (4.918%) | 3 (2.479%) |  |
| PLMI, number of leg movements/h of monitoring time (%) |  |  | 0.8073 |
| None (<15) | 56 (45.902%) | 65 (53.719%) |  |
| Low (15–<45) | 39 (31.967%) | 30 (24.793%) |  |
| High (≥45) | 27 (22.131%) | 26 (21.488%) |  |
| Chronotype variability, mean (SD) | 0.526 (0.263) | 0.658 (0.405) | **0.00299** |
| SD, Standard Deviation; CDR, Clinical Dementia Rating; ApoE4 status = apolipoprotein E ε4 positive ; CSF: cerebrospinal fluid; Aβ, amyloid-beta; AHI, Apnea-Hypopnea Index; PLMI, Periodic Limb Movement Index |  |  |  |

**Table 2:** EEG Sleep Parameters.

| **Table 2: EEG Sleep Parameters** |  |  |  |
| --- | --- | --- | --- |
|  | **Early chronotype** | **Late chronotype** | **p-value** |
|  | Midpoint of sleep before 3:04 AM (n = 122) | Midpoint of sleep after 3:04 AM (n = 121) |  |
| Total sleep time, min, mean (SD) | 371.7555 (57.798) | 377.9669 (66.287) | 0.4452 |
| Sleep efficiency, %, mean (SD) | 78.77617 (9.115) | 78.24584 (9.871) | 0.6693 |
| Sleep onset latency, min, mean (SD) | 17.19489 (11.865) | 19.06408 (13.855) | 0.2684 |
| Wake after sleep onset, min, mean (SD) | 141.076 (21.207) | 144.8782 (29.037) | 0.2454 |
| N2 and N3 time, min, mean (SD) | 254.2989 (51.253) | 270.4797 (63.122) | **0.03231** |
| REM time, min, mean (SD) | 81.13711 (25.163) | 72.3852 (24.22) | **0.007088** |
| REM latency, min, mean (SD) | 99.99076 (62.856) | 128.29124 (78.748) | **0.002652** |
| NREM SWA <1 Hz, $\mu\text{V}^2/\text{Hz}$, mean (SD) | 58.04261 (40.302) | 75.09838 (60.766) | **0.01248** |
| NREM SWA 1-2 Hz, $\mu\text{V}^2/\text{Hz}$, mean (SD) | 39.58187 (25.023) | 49.44853 (33.736) | **0.012** |
| NREM SWA 2-3 Hz, $\mu\text{V}^2/\text{Hz}$, mean (SD) | 12.01186 (6.869) | 13.5532 (7.187) | 0.09507 |
| NREM SWA 3-4 Hz, $\mu\text{V}^2/\text{Hz}$, mean (SD) | 5.332111 (2.825) | 5.830836 (2.917) | 0.1855 |
| Min, Minutes; SD, Standard Deviation; N2, Stage N2 sleep; N3, Stage N3 sleep; REM, Rapid-Eye Movement sleep; NREM SWA, Non-REM Slow Wave Activity; Hz, Hertz |  |  |  |

**Table 3:** Actigraphy and Self-Reported Sleep Parameters.

| **Table 3: Actigraphy and Self-Reported Sleep Parameters** |  |  |  |
| --- | --- | --- | --- |
|  | **Early chronotype** | **Late chronotype** | **p-value** |
|  | Midpoint of sleep before 3:04 AM<br>(n = 122) | Midpoint of sleep after 3:04 AM<br>(n = 121) |  |
| Actigraphic total sleep time, min, mean (SD) | 390.8058 (51.996) | 397.5094 (64.132) | 0.3719 |
| Actigraphic sleep efficiency, %, mean (SD) | 81.65731 (7.291) | 80.40583 (9.184) | 0.2409 |
| Actigraphic sleep onset latency, min, mean (SD) | 19.73729 (15.659) | 24.01861 (24.961) | 0.1122 |
| Actigraphic wake after sleep onset, min, mean (SD) | 149.4696 (21.075) | 153.4047 (29.456) | 0.2327 |
| Actigraphic number of awakenings, n, mean (SD) | 32.65246 (12.499) | 34.74959 (12.87) | 0.1988 |
| Self-reported total sleep time, min, mean (SD) | 427.47408 (62.1) | 446.27718 (69.78) | **0.03165** |
| Self-reported sleep onset latency, min, mean (SD) | 25.52586 (23.949) | 23.94115 (19.837) | 0.5857 |
| Self-reported number of awakenings, n, mean (SD) | 2.126496 (1.143) | 2.086725 (1.471) | 0.8196 |
| Min, minutes; SD, Standard Deviation |  |  |  |

Participants with later chronotypes had lower REM time, longer REM onset latency (Figure 2), and greater time in stages N2 and N3 (Table 2). Self-reported total sleep time, but not objective measures of sleep time measured by either EEG or actigraphy, was significantly longer in those with later chronotypes. <1 Hz and 1-2 Hz NREM slow-wave activity was higher in participants with late chronotype (Figure 3; Table 2). Notably, individuals with later chronotypes also had significantly greater between-night chronotype variability.

**Figure 2:**
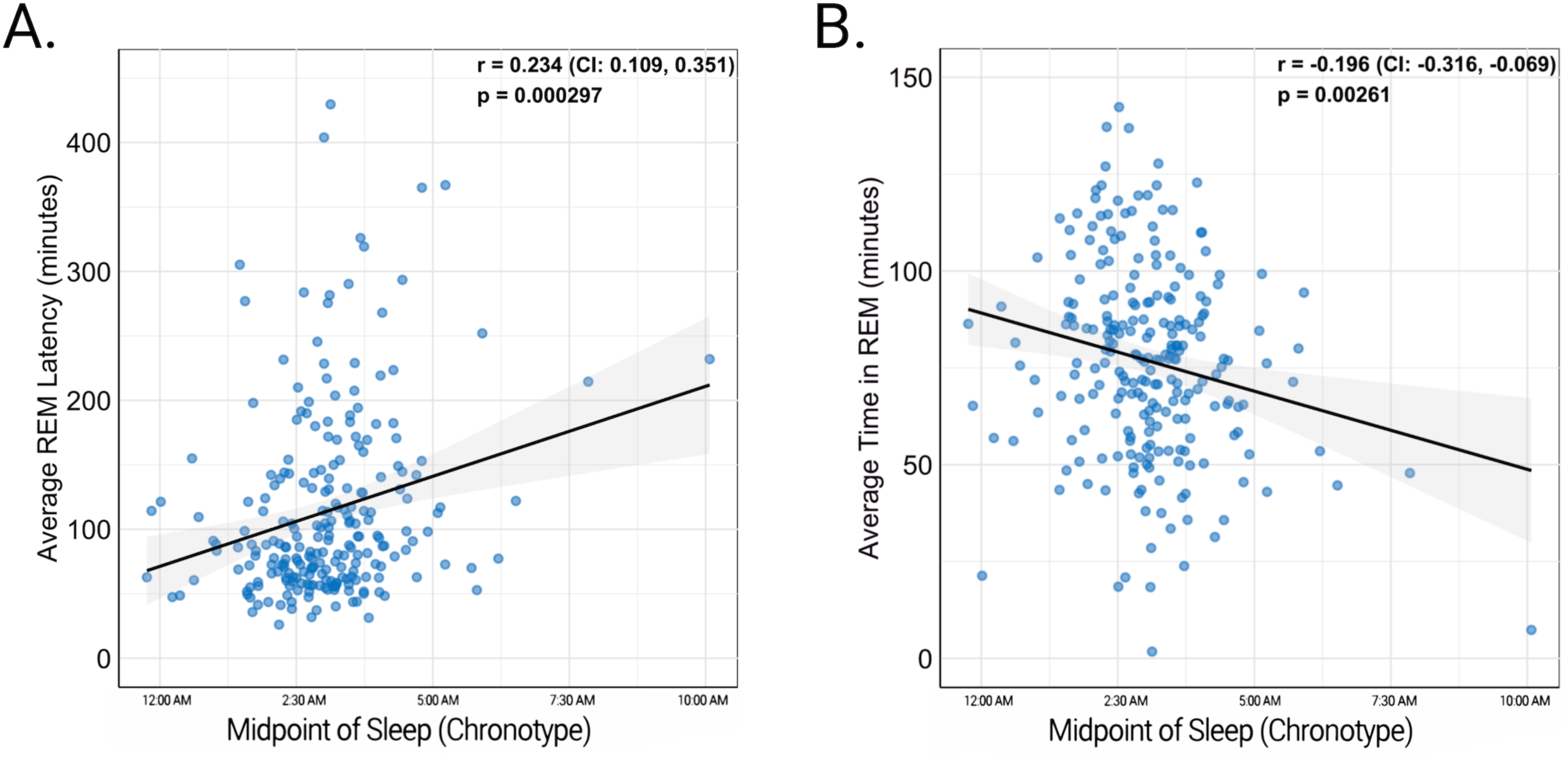
Relationship of midpoint of sleep and REM sleep. As the midpoint of sleep (chronotype) became later in the night, average REM onset latency increased (A) and average time in REM sleep decreased (B).

**Figure 3:**
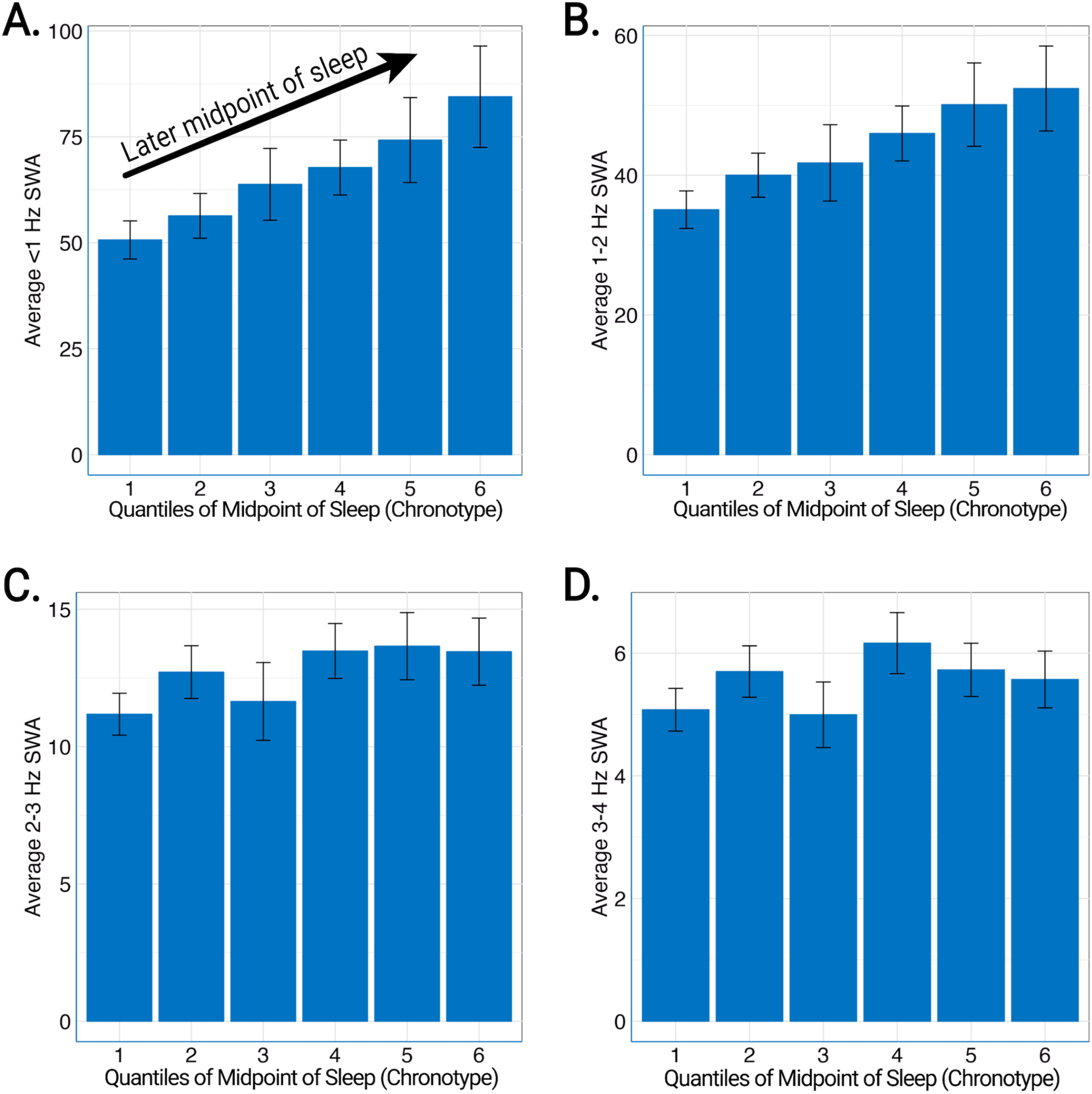
Relationship of midpoint of sleep and NREM slow wave activity. Rounded to the nearest minute, the midpoint of sleep was separated into quantiles of 11:46 PM to 2:07 AM (first, leftmost), 2:07 AM to 2:38 AM (second), 2:38 AM to 3:03 AM (third), 3:03 AM to 3:28 AM (fourth), 3:28 AM to 3:55 AM (fifth), 3:55 AM to 10:04 AM (sixth, rightmost). <1 Hz and 1-2 Hz NREM slow wave activity (SWA) increased with later midpoint of sleep (A, B). The same relationship was not seen for 2-3 Hz NREM SWA (C) or 3-4 Hz (D).

### EEG sleep parameters

Multiple EEG-based sleep parameters have been associated with cognitive deficits and AD pathology. However, sleep may be confounded by factors such as age, sex, ApoE4+ status, amyloid pathology, and others.^53–56^ Therefore, we adjusted for age, sex, race, CDR, ApoE4 status, CSF Aβ42/40 ratio, AHI, and PLMI to assess the relationship between EEG-derived sleep parameters and the midpoint of sleep (chronotype). In the fully adjusted model, later chronotype was associated with longer REM onset latency and shorter time in REM (Table 4). Longer REM onset latency was also associated with female sex, mild cognitive impairment, and ApoE4 status. In contrast, biomarker evidence of amyloid pathology was associated with decreased REM onset latency. Chronotype was not significantly associated with time in NREM stages 2 and 3, total sleep time, sleep efficiency, sleep onset latency, or wake after sleep onset (Table 4, 5).

**Table 4:** EEG Sleep Stages.

|  | Dependent Variables |  |  |  |  |  |
| --- | --- | --- | --- | --- | --- | --- |
|  | Time in N2 and N3 (min) |  | Time in REM (min) |  | REM Onset Latency (min) |  |
| Covariates | Estimate (β) | p-value | Estimate (β) | p-value | Estimate (β) | p-value |
| Chronotype | 4.385 | 0.150 | -3.911 | **0.004** | 12.908 | **<0.001** |
| Age | 0.890 | 0.220 | -0.911 | **0.005** | 0.512 | 0.551 |
| Sex | 21.287 | **0.004** | 2.035 | 0.530 | 47.839 | **<0.001** |
| Race | -29.311 | **0.021** | -9.993 | 0.075 | -25.951 | 0.084 |
| CDR | 9.151 | 0.346 | -4.525 | 0.292 | 40.501 | **<0.001** |
| ApoE4 Status | 11.123 | 0.190 | 1.352 | 0.718 | 21.078 | **0.036** |
| Aβ42/40 | 0.329 | 0.999 | -51.485 | 0.528 | 486.990 | **0.027** |
| AHI | -1.150 | **0.004** | 0.009 | 0.956 | 0.623 | 0.179 |
| PLMI | 0.001 | 0.996 | -0.039 | 0.571 | 0.331 | 0.073 |

Min, Minutes; SD, Standard Deviation; N2, Stage N2 sleep; N3, Stage N3 sleep; REM, Rapid-Eye Movement sleep; CDR, Clinical Dementia Rating; ApoE4 status = apolipoprotein E ε4 positive; Aβ, amyloid-beta; AHI, Apnea-Hypopnea Index; PLMI, Periodic Limb Movement Index

**Table 5:** EEG Sleep Parameters.

|  | Dependent Variables |  |  |  |  |  |  |  |
| --- | --- | --- | --- | --- | --- | --- | --- | --- |
|  | TST (min) |  | Sleep Efficiency (%) |  | SOL (min) |  | WASO (min) |  |
| Covariates | Estimate (β) | p-value | Estimate (β) | p-value | Estimate (β) | p-value | Estimate (β) | p-value |
| Chronotype | 3.754 | 0.257 | -0.790 | 0.128 | 0.761 | 0.278 | 1.792 | 0.191 |
| Age | -0.003 | 0.997 | -0.048 | 0.699 | 0.035 | 0.833 | -0.069 | 0.830 |
| Sex | 23.347 | **0.004** | 0.701 | 0.574 | 6.147 | **<0.001** | 10.142 | **0.002** |
| Race | -41.549 | **0.003** | -3.289 | 0.128 | 1.176 | 0.687 | -4.806 | 0.402 |
| CDR | 11.272 | 0.285 | -0.893 | 0.589 | 1.875 | 0.401 | 4.483 | 0.290 |
| ApoE4 Status | 9.077 | 0.324 | 0.692 | 0.631 | -0.355 | 0.855 | 4.853 | 0.201 |
| Aβ42/40 | -169.20 | 0.399 | -25.405 | 0.419 | 41.176 | 0.332 | -29.508 | 0.720 |
| AHI | -0.377 | 0.375 | -0.155 | **0.020** | 0.045 | 0.619 | -0.138 | 0.427 |
| PLMI | 0.002 | 0.990 | 0.006 | 0.822 | -0.029 | 0.416 | -0.056 | 0.416 |
| TST, Total Sleep Time; SOL, Sleep Onset Latency; WASO, Wake After Sleep Onset; min, minutes; CDR, Clinical Dementia Rating; ApoE4 status = apolipoprotein E ε4 positive; Aβ, amyloid-beta; AHI, Apnea-Hypopnea Index; PLMI, Periodic Limb Movement Index |  |  |  |  |  |  |  |  |

### Non-REM slow-wave activity

The observed difference in NREM SWA between early and late chronotype (Figure 3; Table 2) could be due to other factors such as age, sex, and amyloid pathology. We tested this in the fully adjusted model and found that later chronotype was significantly associated with higher <1 Hz and 1-2 Hz NREM SWA (Table 6). Female sex was associated with higher NREM SWA in all frequency ranges. As previously reported,^57,58^ lower NREM SWA (<1 Hz, 1-2 Hz, 2-3 Hz) was significantly correlated with lower CSF Aβ42/40 ratio (a marker for amyloid deposition). African Americans also showed decreased <3 Hz NREM SWA.

**Table 6:**
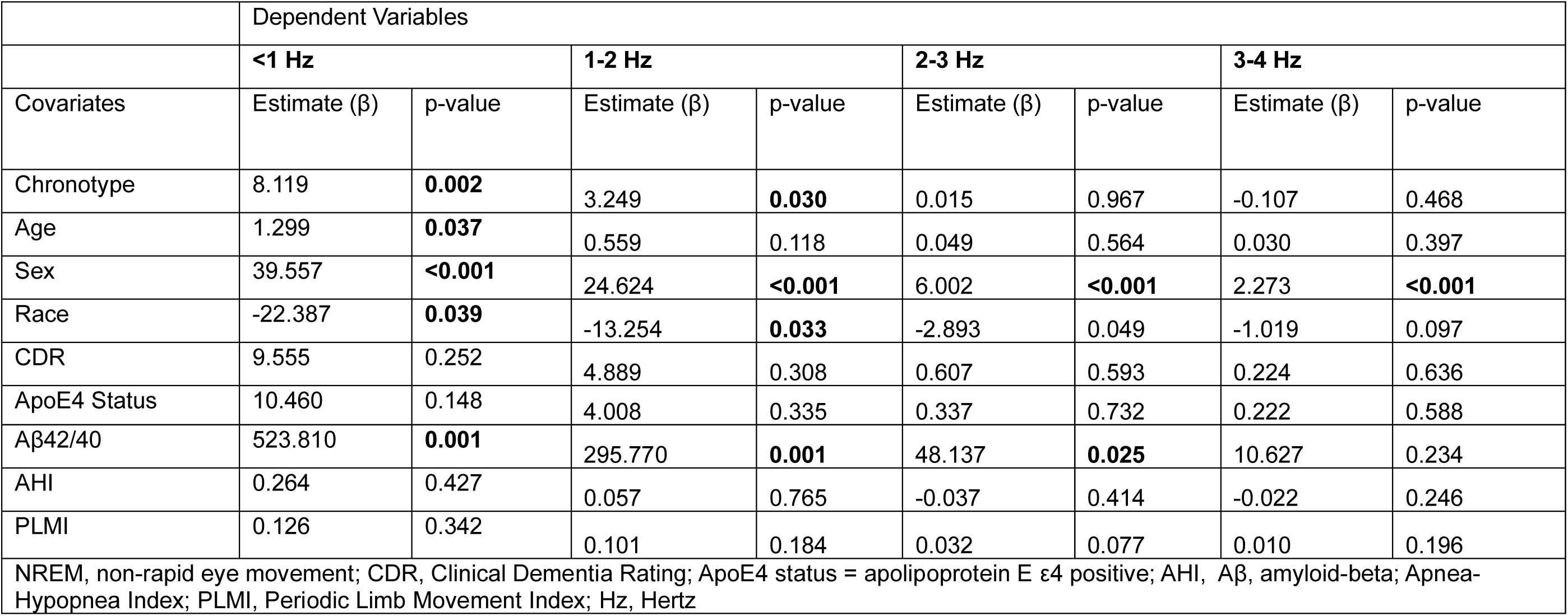
NREM Slow Wave Activity.

|  | Dependent Variables |  |  |  |  |  |  |  |
| --- | --- | --- | --- | --- | --- | --- | --- | --- |
|  | <1 Hz |  | 1-2 Hz |  | 2-3 Hz |  | 3-4 Hz |  |
| Covariates | Estimate (β) | p-value | Estimate (β) | p-value | Estimate (β) | p-value | Estimate (β) | p-value |
| Chronotype | 8.119 | **0.002** | 3.249 | **0.030** | 0.015 | 0.967 | -0.107 | 0.468 |
| Age | 1.299 | **0.037** | 0.559 | 0.118 | 0.049 | 0.564 | 0.030 | 0.397 |
| Sex | 39.557 | **<0.001** | 24.624 | **<0.001** | 6.002 | **<0.001** | 2.273 | **<0.001** |
| Race | -22.387 | **0.039** | -13.254 | **0.033** | -2.893 | 0.049 | -1.019 | 0.097 |
| CDR | 9.555 | 0.252 | 4.889 | 0.308 | 0.607 | 0.593 | 0.224 | 0.636 |
| ApoE4 Status | 10.460 | 0.148 | 4.008 | 0.335 | 0.337 | 0.732 | 0.222 | 0.588 |
| Aβ42/40 | 523.810 | **0.001** | 295.770 | **0.001** | 48.137 | **0.025** | 10.627 | 0.234 |
| AHI | 0.264 | 0.427 | 0.057 | 0.765 | -0.037 | 0.414 | -0.022 | 0.246 |
| PLMI | 0.126 | 0.342 | 0.101 | 0.184 | 0.032 | 0.077 | 0.010 | 0.196 |
| NREM, non-rapid eye movement; CDR, Clinical Dementia Rating; ApoE4 status = apolipoprotein E ε4 positive; AHI, Aβ, amyloid-beta; Apnea-Hypopnea Index; PLMI, Periodic Limb Movement Index; Hz, Hertz |  |  |  |  |  |  |  |  |

### Actigraphy and self-reported sleep parameters

For sleep parameters measured by actigraphy, lower sleep efficiency, longer sleep onset latency, and more awakenings were associated with later chronotype (Table 7). Interestingly, higher CDR and AHI were both significantly associated with lower sleep efficiency, longer sleep onset latency, and a greater number of awakenings showing significance. For self-report sleep parameters, later chronotype was associated with longer total sleep time and higher CDR (Table 8).

**Table 7:** Actigraphy Sleep Variables.

|  | Dependent Variables |  |  |  |  |  |  |  |  |  |
| --- | --- | --- | --- | --- | --- | --- | --- | --- | --- | --- |
|  | TST (min) |  | Sleep Efficiency (%) |  | SOL (min) |  | WASO (min) |  | Awakenings (#) |  |
| Covariates | Estimate (β) | p-value | Estimate (β) | p-value | Estimate (β) | p-value | Estimate (β) | p-value | Estimate (β) | p-value |
| Chronotype | 4.806 | 0.117 | -0.958 | **0.022** | 2.897 | **0.007** | 1.739 | 0.208 | 2.215 | **0.001** |
| Age | 0.423 | 0.557 | -0.050 | 0.611 | -0.036 | 0.888 | -0.032 | 0.920 | 0.054 | 0.723 |
| Sex | 25.581 | **0.001** | 1.527 | 0.127 | -0.393 | 0.879 | 9.470 | **0.005** | -2.460 | 0.111 |
| Race | -32.334 | **0.012** | -2.821 | 0.107 | 2.639 | 0.557 | -4.493 | 0.438 | -0.233 | 0.931 |
| CDR | -16.333 | 0.085 | -6.577 | **<0.001** | 13.740 | **<0.001** | 3.885 | 0.364 | 6.363 | **0.002** |
| ApoE4 Status | 9.144 | 0.281 | 0.057 | 0.960 | 1.729 | 0.561 | 5.244 | 0.171 | -0.637 | 0.720 |
| Aβ42/40 | 65.202 | 0.724 | 7.930 | 0.752 | -61.213 | 0.346 | -32.179 | 0.699 | 4.387 | 0.910 |
| AHI | -0.634 | 0.104 | -0.185 | **0.001** | 0.417 | **0.003** | -0.186 | 0.289 | 0.411 | **<0.001** |
| PLMI | -0.204 | 0.186 | -0.029 | 0.174 | 0.076 | 0.159 | -0.053 | 0.443 | -0.013 | 0.696 |

TST, Total Sleep Time; SOL, Sleep Onset Latency; WASO, Wake After Sleep Onset; min, minutes; CDR, Clinical Dementia Rating; ApoE4 status = apolipoprotein E ε4 positive ; AHI, Apnea-Hypopnea Index; PLMI, Periodic Limb Movement Index; Aβ, amyloid-beta

**Table 8:**
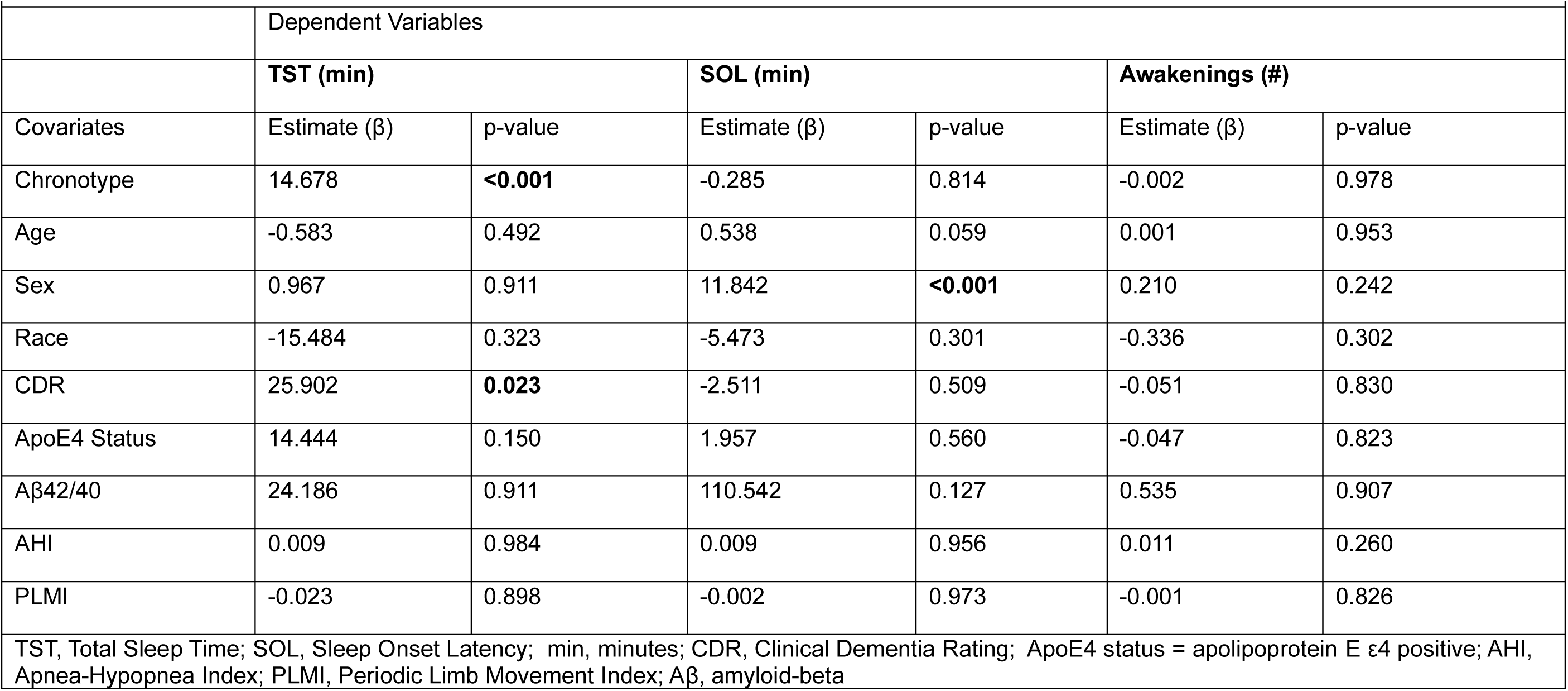
Self-Reported Sleep Variables.

|  | Dependent Variables |  |  |  |  |  |
| --- | --- | --- | --- | --- | --- | --- |
|  | TST (min) |  | SOL (min) |  | Awakenings (#) |  |
| Covariates | Estimate (β) | p-value | Estimate (β) | p-value | Estimate (β) | p-value |
| Chronotype | 14.678 | **<0.001** | -0.285 | 0.814 | -0.002 | 0.978 |
| Age | -0.583 | 0.492 | 0.538 | 0.059 | 0.001 | 0.953 |
| Sex | 0.967 | 0.911 | 11.842 | **<0.001** | 0.210 | 0.242 |
| Race | -15.484 | 0.323 | -5.473 | 0.301 | -0.336 | 0.302 |
| CDR | 25.902 | **0.023** | -2.511 | 0.509 | -0.051 | 0.830 |
| ApoE4 Status | 14.444 | 0.150 | 1.957 | 0.560 | -0.047 | 0.823 |
| Aβ42/40 | 24.186 | 0.911 | 110.542 | 0.127 | 0.535 | 0.907 |
| AHI | 0.009 | 0.984 | 0.009 | 0.956 | 0.011 | 0.260 |
| PLMI | -0.023 | 0.898 | -0.002 | 0.973 | -0.001 | 0.826 |

TST, Total Sleep Time; SOL, Sleep Onset Latency; min, minutes; CDR, Clinical Dementia Rating; ApoE4 status = apolipoprotein E ε4 positive; AHI, Apnea-Hypopnea Index; PLMI, Periodic Limb Movement Index; Aβ, amyloid-beta

## DISCUSSION

In this study, later midpoint of sleep or chronotype was associated with measures reflecting poorer sleep quality such as lower time in REM sleep and longer REM onset latency. This association, however, differed depending on the sleep measurement method used. For instance, a late sleep midpoint was correlated with lower sleep efficiency on actigraphy but not on single-channel EEG. Since a greater number of nighttime awakenings measured by actigraphy were also associated with later midpoint of sleep, this suggests that individuals with later chronotype may have more restless sleep that does not result in wakefulness as recorded on EEG. Similarly, only self-reported total sleep time was related to later chronotype. Total sleep time measured by both actigraphy and single-channel EEG were not correlated with the midpoint of sleep.

Interestingly, <1 Hz and 1-2 Hz NREM SWA increased as the midpoint of sleep was delayed. This association was not observed at faster frequencies of 2-3 Hz and 3-4 Hz, and remained significant even after adjusting for multiple potential factors that affect NREM SWA including age, sex, and amyloid pathology (i.e., CSF Aβ42/40). Since individuals with later chronotype had evidence of poorer sleep quality (e.g., less time in REM sleep, lower sleep efficiency, greater nighttime awakenings), we hypothesize that higher NREM SWA may represent a homeostatic response to decreased sleep quality.

Individuals with a self-reported Black ethnic background have been reported to have earlier chronotypes than individuals of self-reported White ethnic background^59^ and African Americans have been reported to have a shorter circadian period than European Americans.^60^ In contrast, African American participants in our cohort had later chronotypes. For example, a study from the UK Biobank cohort found that a matched sample (N=2044; 50% self-reported Black ethnic background) with a mean age of 52 had a significantly greater prevalence of self-reported morning types based on a single question survey item in participants with self-reported Black ethnic background.^59^ The current contrary finding could be due to the comparatively smaller sample size (9.9% African American in the current cohort), the increased age of participants in the present study (mean age = 74), a cohort effect such as geographical location or background characteristics, differences in factors included in each study’s respective models (i.e., AD biomarkers such as CSF Aβ42/40), or methodological differences in identifying chronotype. The participants in the regression analyses may not be representative of the general population, with an educational attainment of 15.3 years among African Americans and 16.5 years among non-African Americans. Although only 24 African American participants completed a lumbar puncture and were included in the fully adjusted models, 63 participants had data available to calculate the midpoint of sleep and the same relationship was observed without adjusting for other factors.

These results indicate that midpoint of sleep or chronotype is associated with measures indicating poorer sleep quality. Since we calculated midpoint of sleep based on each individual’s sleep behavior, we propose that this may be modified as part of a treatment plan to improve sleep quality. Chronotype can be modified by light exposure;^61^ for example, exposure to 9,500 lux resulted in a shift of 4.5 hours earlier in young men.^62^ Melatonin administration resulted in a shift 1.5 hours earlier.^63^ Among subjects with delayed sleep phase syndrome, earlier sleep and wake times coupled with an attempt to avoid nighttime light exposure as well as napping resulted in a shift in chronotype of over 1 hour earlier in the absence of bright light therapy.^64^

CDR was significantly associated with multiple sleep variables, such as REM latency, sleep efficiency, sleep onset latency, awakenings as measured by actigraphy, and self-reported total sleep time. All of these variables were also significantly associated with chronotype. Further, each variable had the same direction of effect with CDR as it did with chronotype. That is, later chronotype and mild cognitive impairment are associated with overlapping aspects of sleep in similar ways. This suggests that the association of later chronotype and poor sleep quality may be related to cognitive dysfunction. Future studies are needed to assess if interventions to advance circadian phase improve measures of sleep quality and cognitive performance and/or behaviors associated with symptomatic AD such as the later-day agitation that is often referred to as sundowning.

Future research is needed to determine if an intervention to normalize very late chronotype results in improved sleep parameters such as REM sleep or decreased intra-night sleep/wake time variability. This could be accomplished through behavioral recommendations and education about the effects of abnormal sleep phase, bright light therapy beginning in the morning, limiting light exposure in the evening, modifying other zeitgebers such as exercise and meal timing, melatonin, or a combination thereof. Further research can also elucidate how inter-night variability in chronotype affects sleep quality and dementia, the interaction between chronotype and cognitive function in sleep outcomes, and the longitudinal association between chronotype and Alzheimer’s disease biomarkers.

This study provides a detailed characterization of the association of the midpoint of sleep or chronotype with a range of sleep parameters in older adults with and without Alzheimer’s disease symptomatology. Results indicate that a later midpoint of sleep is associated with different sleep parameters even after adjustment for potential confounding factors. Several of the sleep measures suggest that worse sleep quality (e.g., decreased time in REM sleep) is associated with later chronotype. However, the relationship of other sleep parameters is unclear and warrants further investigation. For instance, later chronotype is associated with higher NREM SWA. This may be due to an increased homeostatic response to poor sleep quality. NREM SWA has been proposed as a marker of brain function in individuals at risk for AD. Further research is needed to establish the role of chronotype on NREM SWA and other sleep measures and if chronotype should be taken into account when assessing the effect of sleep on AD.

## ACKNOWLEDGEMENTS

We are indebted to the participants for their contributions to the study

## AUTHOR CONTRIBUTIONS

S.S.: Conception and design of the study; acquisition and analysis of data; drafted manuscript and figures; critically reviewed and approved the manuscript for publication.

C.D.T.: Acquisition and analysis of data; critically reviewed and approved the manuscript for publication.

R.R.: Acquisition and analysis of data; critically reviewed and approved the manuscript for publication.

A.P.S.: Critically reviewed and approved the manuscript for publication. J.C.M.: Critically reviewed and approved the manuscript for publication. D.M.H.: Critically reviewed and approved the manuscript for publication.

B.P.L.: Conception and design of the study; acquisition and analysis of data; drafted manuscript and figures; critically reviewed and approved the manuscript for publication.

