## Supplementary figures and images for "Chronotype is Associated with Sleep Quality in Older Adults"

### Supplementary Figure 1

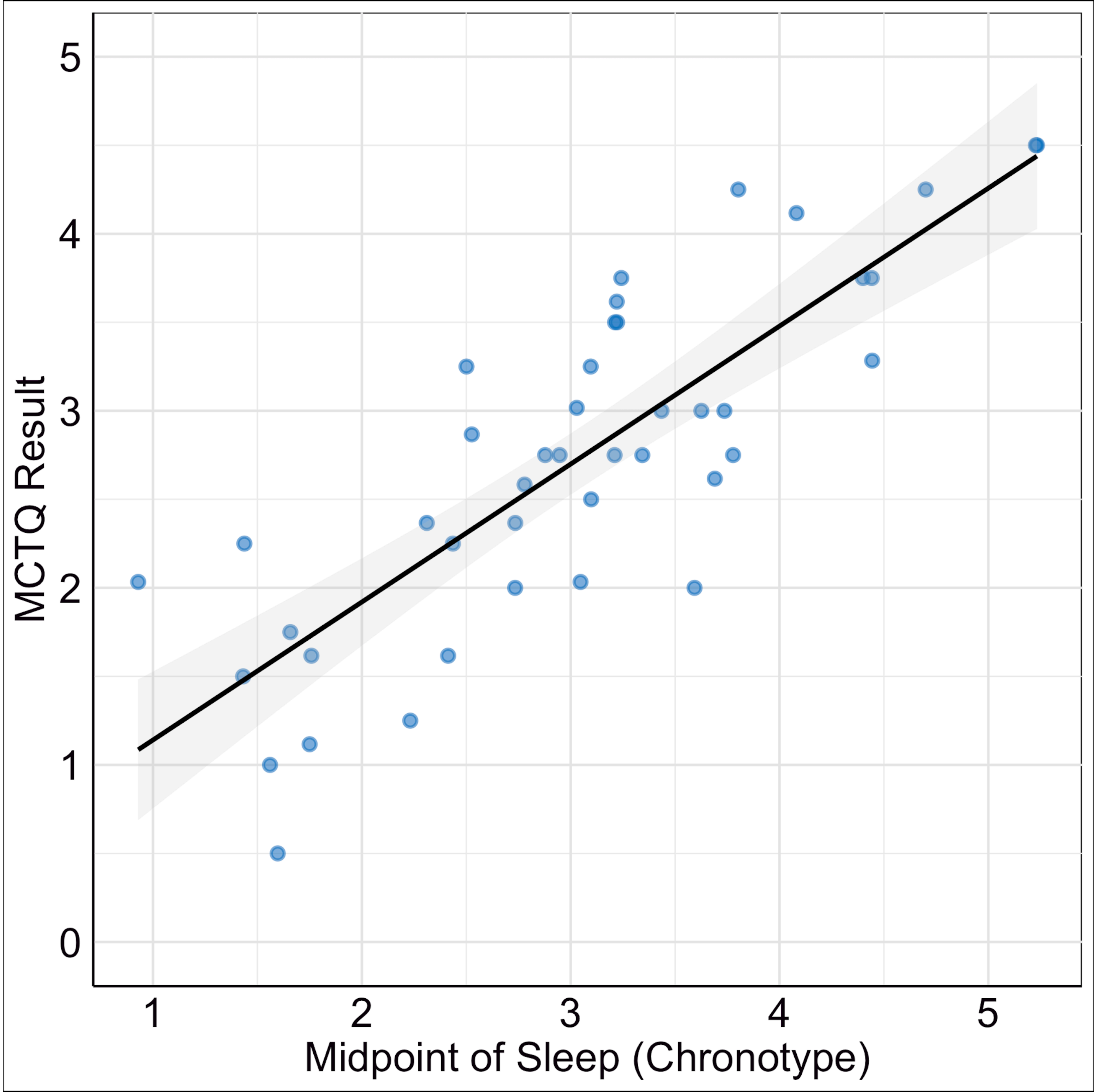
